# Follow the Money: A Closer Look at US Tobacco Industry Marketing Expenditures

**DOI:** 10.1101/2021.08.08.21261761

**Authors:** David T. Levy, Alex Liber, Christopher J. Cadham, Luz María Sánchez-Romero, Andrew Hyland, K. Michael Cummings, Clifford E. Douglas, Rafael Meza, Lisa Henriksen

**Affiliations:** Georgetown University-Lombardi Comprehensive Cancer Center, Cancer Prevention and Control Program, 3300 Whitehaven St. NW, Washington, DC, USA; Department of Health Management and Policy, University of Michigan School of Public Health, 1415 Washington Heights, Ann Arbor, MI, USA; Department of Health Behavior, Roswell Park Comprehensive Cancer Center, Buffalo, NY, USA; Medical University of South Carolina, Department of Psychiatry & Behavioral Sciences, Charleston, SC, USA; Department of Epidemiology, University of Michigan School of Public Health, 1415 Washington Heights, Ann Arbor, MI, USA; Stanford Prevention Research Center, Stanford University School of Medicine, Palo Alto, CA, USA

**Author notes:** Corresponding Author: David T. Levy, PhD, Cancer Prevention and Control, Lombardi Comprehensive Cancer Center, Georgetown University, 3300 Whitehaven St., NW, Suite 4100, Washington, DC 20009.

## Abstract

**Introduction:** While much of the concern with tobacco industry marketing has focused on direct media advertising, a less explored form of marketing strategy is to discount prices. Price discounting is important because it keeps the purchase price low and can undermine the impact of tax increases.

**Methods:** We examine annual marketing expenditures from 1975 to 2019 by the largest cigarette and smokeless tobacco companies. We consider three categories: direct advertising, promotional allowances, and price discounting. In addition to considering trends in these expenditures, we examine how price discounting expenditures relate to changes in product prices and excise taxes.

**Results:** US direct advertising expenditures for cigarettes fell from 80% of total industry marketing expenditures in 1975 to less than 3% in 2019, while falling from 39% in 1985 to 6% in 2019 for smokeless tobacco. Price-discounting expenditures for cigarettes became prominent after the Master Settlement Agreement and related tax increases in 2002. By 2019, 87% of cigarette marketing expenditures were for price discounts and 7% for promotional allowances. Smokeless marketing expenditures were similar: 72% for price promotions and 13% for promotional allowances. Price discounting increased with prices and taxes until reaching their currently high levels.

**Conclusions:** While much attention focuses on direct advertising, other marketing practices, especially price discounting, has received less attention. Local, state and federal policies that use non-tax mechanisms to increase tobacco prices and restrict industry contracts with retailers are needed to offset/disrupt industry marketing expenditures. Further study is needed to better understand industry decisions about marketing expenditures.

**Key points:**

- While much of the concern with tobacco industry marketing has focused on direct media advertising, a less explored form of marketing strategy is to discount prices. Price discounting is important because it keeps the purchase price low and can undermine the impact of tax increases, contributing to tobacco initiation and exacerbating socio-economic health disparities.
- While cigarette and smokeless tobacco industry direct marketing expenditures have drastically fallen over time, price-discounting expenditures have dramatically increased in line with increases in prices and taxes.
- Local, state and federal policies that restrict non-tax mechanisms to increase tobacco prices and restrict industry contracts with retailers are needed to offset/disrupt industry marketing expenditures.

## INTRODUCTION

In the 1980s, famed American investor Warren Buffet said, “I’ll tell you why I like the cigarette business… It costs a penny to make. Sell it for a dollar. It’s addictive. And there’s fantastic brand loyalty.” ^1^ While the retail price of cigarettes has gone up, the relative magnitude between manufacturer’s cost of production and marketing, tax burden, and retail price has not markedly changed.^2^

The large profits achieved by cigarette companies are the result of decades of strategic marketing activities, designed to increase and maintain consumer demand for their products. Much of tobacco control policy and research has focused on direct advertising, commonly occurring through various media such as television, radio, magazines, billboards, point of sale, and now social media.^3^ Empirical studies of marketing restrictions often focus only on the impact of direct advertising.^3–8^ Researchers and policymakers have devoted comparatively less attention to indirect marketing, such as eliminating sponsorships, branding, price promotions, and free samples.

A parallel literature focuses primarily on cigarette price promotions. Industry documents and studies^9–11^ have identified five types of price discounting practices: 1) couponing, whereby a consumer is provided a voucher that may be used to directly reduce the price of a tobacco product, 2) free samples, 3) quantity discounts (e.g., lower prices per pack when more than one pack is purchased), 4) reducing the price of brands used by more price-sensitive consumers (such as youth and those of low SES), and 5) geographically targeting price sensitive, less mobile customers in particular areas (e.g., poorer neighborhoods or near schools). A recent study found that, between 2011-2016, 11.3% of cigarettes, 3.4% of large cigars, 4.1% of little cigars, and 3.9% of cigarillo sales were price discounted, with top-selling tobacco brands accounting for 36% of cigarette-discounted sales.^12^ In a representative sample of US tobacco retailers, 75.1% advertised price promotions on tobacco products in 2012, and among cigarette packs purchased from those stores, 31.7% of Marlboro and 14.7% of Newport packs included promotional offers.^13^

Economic analysis shows that firms may price discriminate to increase overall consumer demand as well as profits.^14–16^ Prices are reduced to the most price-sensitive consumers, such as youth and those who are poor,^10,17–19^ thereby increasing tobacco use and disparities. At the same time, prices are increased to less price-sensitive consumers, thereby increasing profits from this group.^20^ Studies provide evidence consistent with this practice. A systematic review found that cigarettes sell for lower prices in areas with lower socioeconomic status populations and higher numbers of young people.^21^ Another study found lower Newport menthol cigarette prices in neighborhoods with the highest quartiles of youth, Black residents, and lower-income households.^22^ Studies also show the importance of couponing as another tool for price discrimination via price discounting.^23–26^ This literature finds that couponing has targeted young adults and those of low education,^24,27–29^ and is associated with increased smoking initiation and reduced cessation.^30–35^ Tobacco control researchers have called for a greater focus on retail settings and especially the role of price discrimination in the marketing of tobacco products.^3,36–38^

While attention has been given to price discounting, the role of price discounting may take on added importance during periods of rapid tax or price increase. In particular, when tax increases are imposed, firms may use price discrimination to blunt the intended impact to reduce smoking.^10,18,39–42^ Since raising tobacco taxes is a particularly effective tobacco control strategy^43^ through its ability to increase prices,^44–46^ price-reducing strategies may have critical implications by shifting less of the tax to those who are more price sensitive, thereby reducing the likelihood that they would not smoke.

The purpose of this paper is to examine trends in the different types of tobacco industry marketing expenditures, including those for price discounting. We analyze marketing expenditures data reported to the Federal Trade Commission (FTC) by the largest US cigarette and smokeless tobacco companies. We focus on price discounting expenditures, which have received less attention from tobacco control researchers,^44,46,47^ but are now far greater than direct advertising expenditures.^3–8^ In addition to considering trends in marketing expenditures, we consider how industry price discounting expenditures are related to the Master Settlement Agreement (MSA) and consumer tobacco product prices and cigarette excise taxes.

## METHODS

We collected marketing expenditure data from the 2019 FTC Cigarette Report and Smokeless Tobacco Report.^48,49^ The FTC collects data on advertising and promotion practices from major companies under a compulsory process. In 2019, cigarette firms included Altria, ITG Holdings USA, Reynolds American, and the Vector Group and smokeless tobacco firms included Altria, North Atlantic Trading Company, Reynolds American, Swedish Match, and Swisher. Marketing expenditures data are provided at the industry level from 1975-2019 for cigarettes and 1985-2019 for smokeless tobacco.

For both cigarettes and smokeless tobacco, we categorize marketing expenditures into three groups: direct advertising, promotional allowances, and price discounts. Direct advertising includes traditional forms of advertising, such as television, radio, newspapers, billboards, and (retail) point-of-sale, as well as direct mail, company website, internet, telephone, social media endorsements, and other advertising and merchandising. Promotional allowances include allowances paid to cigarette retailers and wholesalers to facilitate the sale or placement of a cigarette brand, including payments for volume rebates, incentive payments, value-added services, promotional execution, and satisfaction of reporting requirements (considered promotional allowances), and quantity promotions (e.g., buy two packs, get one free) which may be bundled with the purchased cigarettes (retail value-added). Many of these payments effectively reduce product prices (e.g., by providing customers with multi-pack discounts or by creating incentives for particular retailers to obtain volume rebates by reducing the price to increase sales).^50–52^ Price discounting payments include direct payments to cigarette retailers or wholesalers to reduce the price of cigarettes to consumers, including off-invoice discounts (for selling a specific quantity or a minimum quantity of a given product over a period of time), buy-downs (in which the dealer receives a per-unit payment for agreeing to sell certain units at a discounted price), voluntary price reductions, and trade programs.

We also compare price discounting expenditures to changes in tobacco products prices and taxes. For cigarettes, we obtained average yearly retail prices (including generics) and tax data from the Tax Burden on Tobacco.^2^ These apply uniform practices and are available for the full period of this study. For smokeless tobacco, taxes take multiple forms and are more difficult to calculate, but price measures were developed using data from the FTC Smokeless Tobacco Report by summing the per-unit prices (sales/units) for each category of smokeless tobacco (chew, pouches, etc.) weighted by each category’s sales. To correct for price inflation, prices and taxes were inflated to 2019 dollars using the consumer price index.^53^ To gauge the relative importance of the three categories of marketing expenditures as separate from the impact of expected sales, we compared prices and taxes to price discounting and promotional allowances as a percent of marketing expenditures.

## RESULTS

### Cigarette Marketing Expenditures

Figure 1 shows the time trend of cigarette marketing expenditures in constant (inflation-adjusted) 2019 dollars, with select years provided in Table 1. Overall marketing expenditures increased until 2003 and then began to decline, while the composition of these expenditures has dramatically changed. In 1975, 80% of marketing expenditures were for direct advertising, with about 15% for promotion allowances and 5% for price discounting. Direct advertising constituted the largest percentage of expenditures through 1991 and fell to 3.5% of expenditures by 2019. Retail-value-added and promotions increased through 2002, especially after 1998, when the MSA restricted many forms of direct advertising and eliminated cigarette-branded merchandise.^54^ However, these expenditures began tapering off in 2003, and retail value-added was no longer traced separately from other spending by 2009. Since 2002, at least 68% of annual marketing expenditures were for price discounting and 26% for promotions, appearing to replace the waning retail-value-added expenditures. Promotional allowances further declined in importance, as price discounting expenditures, including free samples and couponing, increased to about 90% of expenditures in 2002. The US Food and Drug Administration (FDA) prohibited free samples in 2017, but some states passed such restrictions before the federal regulation.

**Table 1.** Percent of Direct Advertising, Promotional, Price Discounting, and Total Cigarette and Smokeless Tobacco Marketing Expenditures, Select Years.

| Categories<br>Years | Cigarettes |  |  |  |  | Smokeless Tobacco |  |  |  |  | Source<br>of<br>data:<br>FDA<br>Cigarette<br>Report,<br>FDA |
| --- | --- | --- | --- | --- | --- | --- | --- | --- | --- | --- | --- |
|  | 1975 | 1986 | 1998 | 2003 | 2019 | 1986 | 1998 | 2002 | 2016 | 2019 |  |
| **Direct advertising (%)<sup>1</sup>** | 80.4% | 69.4% | 26.0% | 7.2% | 3.5% | 71.4% | 62.5% | 27.1% | 19.1% | 15.1% |  |
| **Promotional allowances (%)<sup>2</sup>** | 14.7% | 26.4% | 64.7% | 17.2% | 6.7% | 10.7% | 18.0% | 14.6% | 10.9% | 12.6% |  |
| **Price discounting (%)<sup>3</sup>** | 4.9% | 4.1% | 9.3% | 75.6% | 89.8% | 17.9% | 19.5% | 58.3% | 70.0% | 72.3% |  |
| **Total Marketing Expenditures**<br>**(in million 2019\$)** | \$2,334 | \$5,558 | \$10,758 | \$17,795 | \$7,624 | \$187 | \$236 | \$336 | \$792 | \$659 | |

Smokeless Tobacco Report
1. Direct advertising includes newspapers, magazines, outdoor, direct mail, point of sale, sponsorships endorsements, company website, internet, telephone, audio-visual, social media, and other advertising and merchandising. 2. Promotional allowances (also called retail value-added) includes allowances paid to cigarette wholesalers or retailers to facilitate the sale or placement of any cigarette, including payments for volume rebates, incentive payments, value-added services, promotional execution, and satisfaction of reporting requirements and promotions involving free cigarettes (*e.g.*, buy two packs, get one free), whether or not the free cigarettes are physically bundled together with the purchased cigarettes. 3. Price discounts include payments paid to cigarette retailers or wholesalers to reduce the price of cigarettes to consumers, including off-invoice discounts, buy-downs, voluntary price reductions, and trade programs.

**Figure 1.**
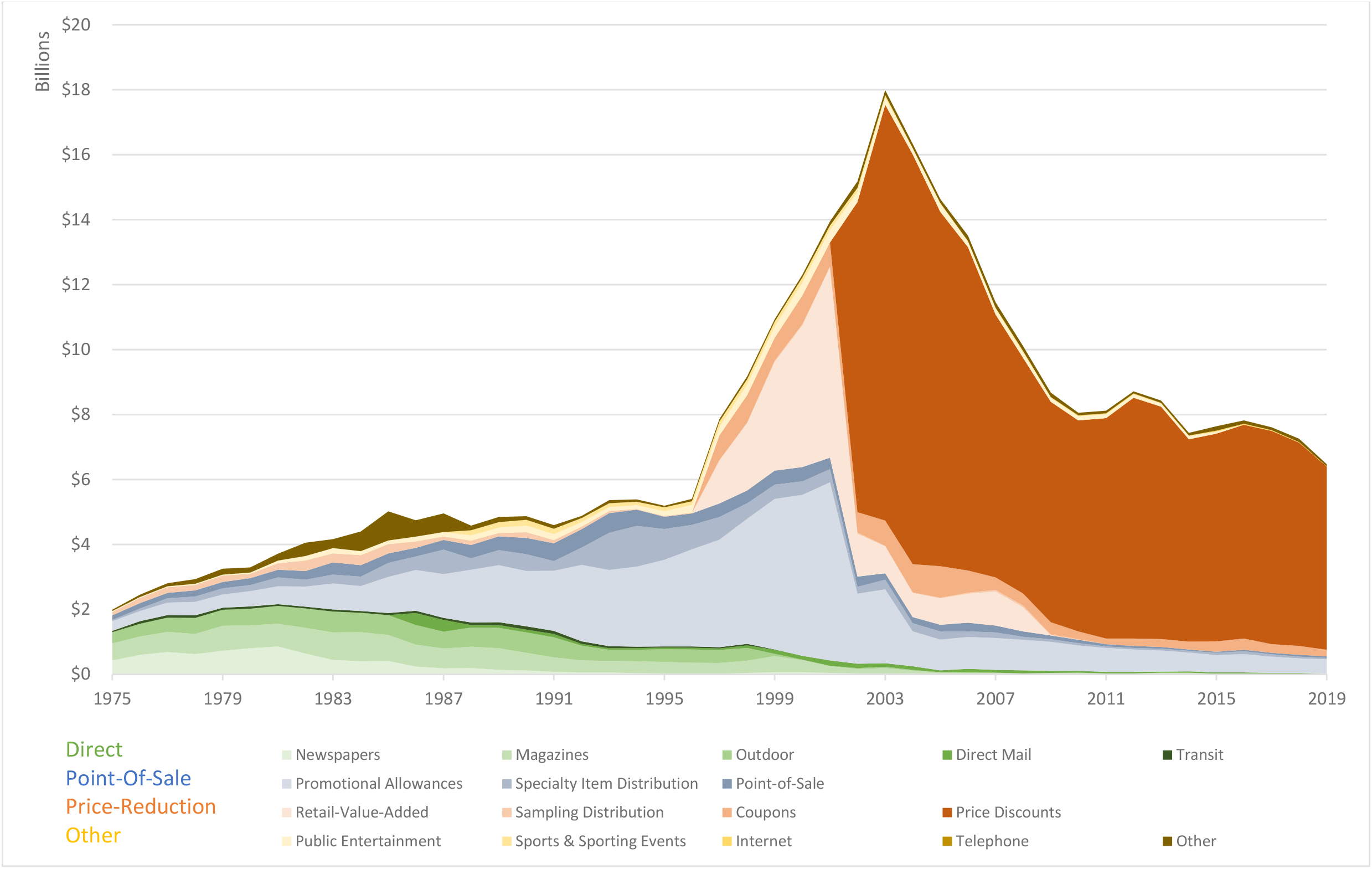
Share of Real US Cigarette Marketing Expenditure (Federal Trade Commission, 1975-2019)

In 2019, the major cigarette manufacturers spent $7.6 billion on cigarette marketing. Of this, $6.6 billion (86.7% of total marketing expenditures, of which 74.7% went to retailers and 12.0% to wholesalers) were for price discounting. Coupons constituted an additional 3% of total expenditures. Spending on promotional allowances in 2019 was 6.7% of marketing expenditures. When combined with price discounts, these indirect marketing efforts accounted for 96.5% of all marketing expenditures. The remaining 3.5% of expenditures in 2019 was for direct advertising, including 0.8% for point-of-sale, 0.1% for magazines, 0.3% for direct mail, 0.7% for specialty item distribution, 0.6% for adult-only public entertainment, 0.2% for company websites and 0.9% for “other.”

### Cigarette Marketing Expenditures in Relationship to Taxes and Price

Figure 2 shows that the percent price discounting and promotional allowances began increasing in about 1980, although mostly due to increased promotional allowances. Larger percentage increases in these expenditures occurred after 1997, when the MSA was implemented and cigarette tax increases became more prevalent. Figure 2 also shows a similar, but more direct relationship of these expenditures to cigarette prices through 2003. The percent of price discounting and promotional expenditures increased only slightly in 2009 when there was a large federal tax increase and prices continued to increase. However, at this point, price discounting expenditures had reached about 93% of total marketing expenses and showed minimal increase with prices or taxes going forward.

**Figure 2.**
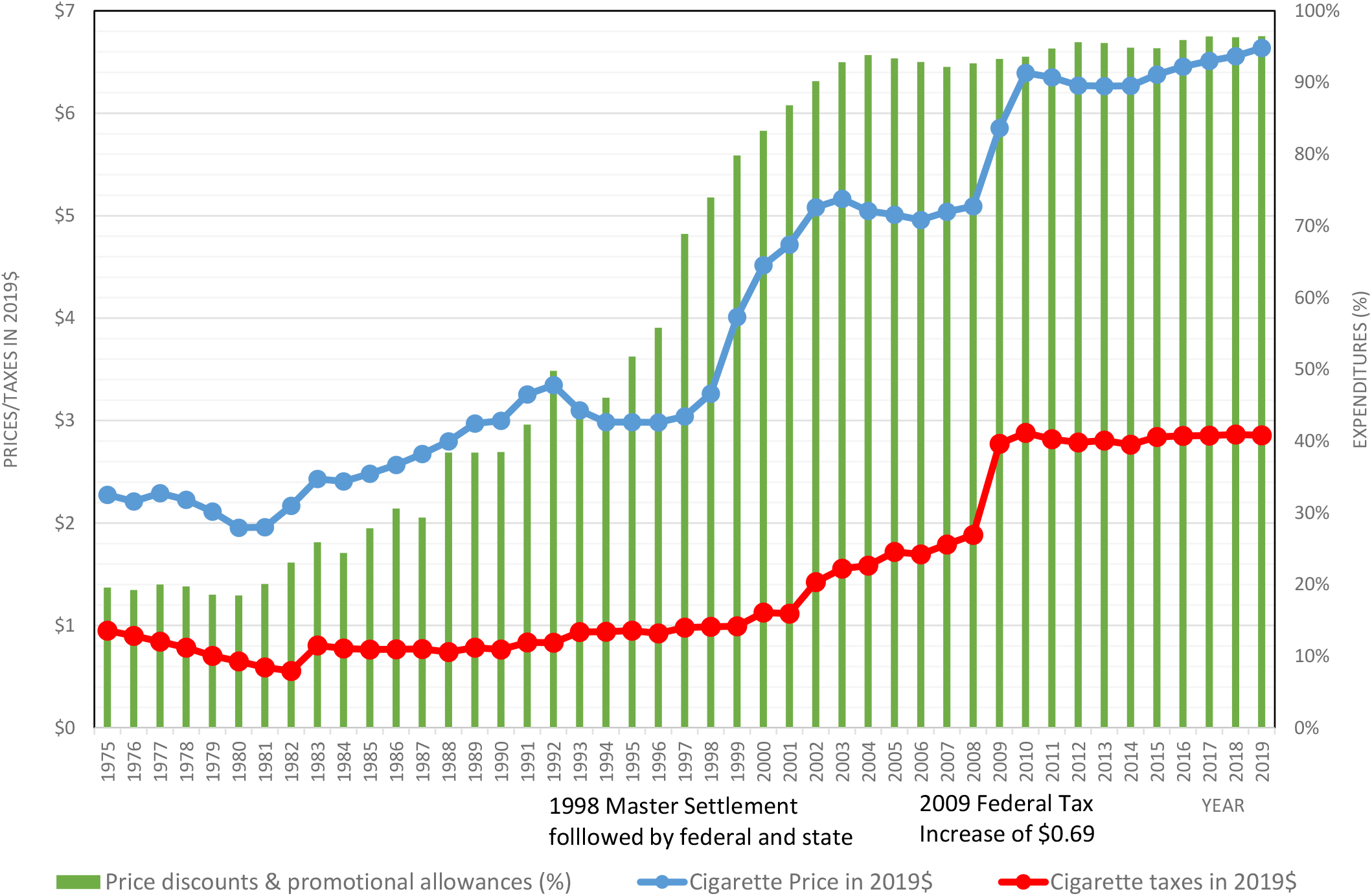
Price Discounts and Promotional Allowances as Percent of Marketing Expenditures in Billion 2019$ vs. Cigarette Prices and Excise Taxes in 2019$, 1975-2019.

### Smokeless Tobacco Marketing Expenditures

As shown in Table 1, smokeless tobacco total marketing expenditures in 2019 dollars peaked in 2016, and then fell. In 1986, direct advertising promotions (mostly magazines, point-of-sale and outdoors after 1986) were 71% of expenditures and have since steadily declined to 15% in 2019. Starting in 1998, expenditures began shifting rapidly to promotions and retail value-added. Coupons and sampling were important in 1998, but sampling began declining in 2006 and coupons have fluctuated between 6% and 14% of expenditures since 1998. In 2002, price discounting became a separate and important component (58%) of overall marketing expenditures, with the largest increases in 2006 and 2013. Promotional allowances have been at least 10% of expenditures since 2009.

In 2019, the major smokeless tobacco manufacturers spent $576 million on marketing expenditures. The companies reported 65.3% of total marketing expenditures to price discounting, with an additional 0.1% on sampling and 6.9% on coupons. Manufacturer spending on promotional allowances was 12.6%. Direct advertising was 15.1% of total marketing expenditures, mostly point-of--sale (3.4%), magazines (1.1%), direct-mail (0.6%), company websites (0.9%), non-branded specialty-item distribution (2.4%) and consumer engagement in adult-only facilities (0.9%).

### Smokeless Tobacco Marketing Relative to Price

Figure 3 shows that the percent of price discounting and promotional expenditures increased slowly with inflation-adjusted smokeless tobacco prices mostly just before 1997, but then rapidly increased in 1998. After prices and expenditures fell between 2003 to 2008, prices and price discounting and promotional allowances both began increasing in 2008.

**Figure 3.**
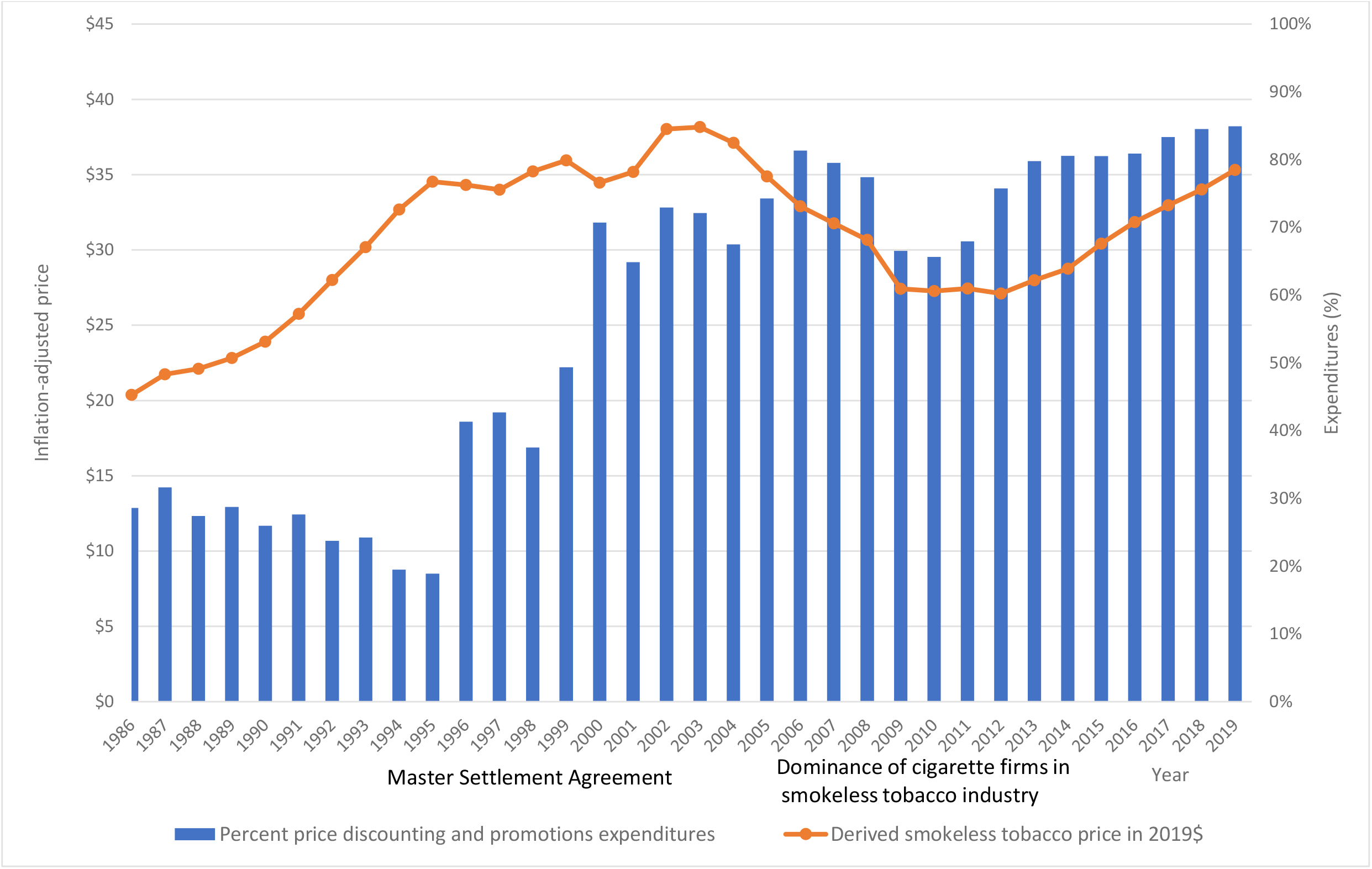
Price Discounts and Promotional Allowances as Percent of Marketing Expenditures in Billion 2019$ vs. Smokeless Tobacco Prices in 2019$, 1986-2019.

## DISCUSSION

Our study considers the relative importance of various marketing strategies and changes over time. While direct advertising has been a major focus of tobacco control research, the majority of US marketing expenditures over the last 20 years were related to price discounting and related promotions. Annual expenditures on direct advertising for cigarettes fell from 80% of the industry’s marketing expenditures in 1975 to 3.5% in 2019, while direct advertising for smokeless tobacco fell from 71% in 1985 to 15% in 2019. Meanwhile, price-discounting promotion expenditures for cigarettes increased dramatically since 1997, accounting for almost 90% of outlays by 2019. Another 7% of 2019 expenditures went towards promotional allowances, which also contributes to reduced prices. For smokeless tobacco, 72% of marketing expenditures were for price discounting with another 13% for promotional allowances in 2019.

The data shows a clear relationship of discounting practices to retail prices and the MSA through 2003. As in previous studies,^55,56^ the trend away from direct advertising and towards price-discounting and promotional expenditures began accelerating after 1997 when the MSA restricted cartoons, transit advertising, most outdoor advertising, product placement in media, branded merchandise, free samples (except in adult-only facilities), and most sponsorships.^54^ Around the same time, federal taxes increased from $0.24 to $0.34 per pack on January 1, 2000, and then to $0.39 per pack on January 1, 2002, and state taxes were also increasing.^6^ In addition, other tobacco control policies (e.g., smoke-free air laws and media campaigns) were ramping up in many states.^57–60^ Cigarette price discounting expenditures increased to 78% by 2009 and 85% by 2013, with a $0.62 federal tax increase and the Family Smoking Prevention and Tobacco Control Act (FSPTCA) in 2009.^57^ The percent devoted to price discounting and promotion allowances, together, reached 93% of total marketing expenses in 2002 and then showed minimal increases over the next 17 years, peaking at 96.5% in 2019, perhaps suggesting that price discounting and promotional allowances had reached a saturation point as prices or taxes continued to increase. For smokeless tobacco, price-discounting expenditures showed substantial increases in about 1998 and then again in 2008, not long after Reynolds American acquired Conwood Smokeless Tobacco Company (2006) and introduced Camel Snus and shortly before Altria acquired the U.S. Smokeless Tobacco Company (2009).^62,63^ While we suggest how patterns of marketing expenditures are related to taxes and other tobacco control policies, further exploration of the timing of these changes is warranted.

Our analysis focuses price-related expenditures as a percent of total marketing expenditures in order to focus on the relative importance of price discounting in marketing. However, total marketing expenditures patterns have also changed over time. Cigarette marketing expenditures increased through 2003. Since 2003, total marketing expenditures fell less than proportionally to pack sales to half their 2003 level (Supplement Material available from author),^64^ while total price discounts and promotional allowance expenditures fell in proportion to pack sales (Supplement Material available from author). These reductions in marketing expenditures may reflect MSA restrictions, but may also reflect the generally declining cigarette sales resulting from stronger tobacco control policies, and shifts toward little cigars, smokeless tobacco, and e-cigarettes.^65^ In contrast, smokeless tobacco price discounting expenditures have risen sharply, particularly since the cigarette companies started dominating the industry in 2005 (Supplement Material available from author). The role of absolute versus percent price discounting expenditures warrants further study.

A useful framework for understanding the importance of these marketing practices is the 4Ps: Promotions, Price, Product, and Place.^3^ While the role of Promotion, commonly understood as direct advertising, and its impact on tobacco use is well-documented, US tobacco companies have increasingly shifted their marketing emphasis to Price. This focus has important implications since taxes are an important tobacco control policy and discounting offsets some of the effects of tax-related price increases.^44–46^ In particular, cigarette companies direct these discounts to those who are more price sensitive, youth and young adults and to those of lower SES,.^16,19,44,46,47,66,67^ Thereby, discounts can encourage initiation among youth and discourage cessation among users who are young adults or economically disadvantaged. Studies also indicate that discounts tend to be received by high-intensity smokers, who are more price-sensitive lower-intensity smokers.^30,66,68–70^ Thereby, price discounting may also discourage cessation and increase the quantity smoked among those at highest health risk.^71^ Further study of price discounting and the relationship of prices to taxes would improve understanding of the simultaneous over-shifting and under-shifting of taxes.

To gauge the potential importance of price discounting, we calculated the potential role of related expenditures relative to prices. Dividing price discounting and promotional expenditures ($7.35 billion) by the number of packs (cigarette-tax paid stick sales (202.9 billion)^64^/20), we calculated that price discounting expenditures translate to about $0.73 per pack. With prices per pack estimated at $7.22 in 2021,^72^ the average per-pack discount translates to an average price reduction of ∼10%. With an overall smoking prevalence price elasticity of -0.3%,^44^ a 10% price reduction uniformly applied to all customers would have kept smoking prevalence 3% higher than if not applied. However, these industry expenditures will likely to lead to much greater impact, since the discounts are applied to more price-sensitive customers (e.g., youth, low income and more frequent users). Although slightly offset by price increases to less price-sensitive consumers, smoking prevalence would be effectively increased by much more than 3%. In addition, price discounts may be exacerbated if retail and wholesale firms reduce the price more than proportionately to marketing expenditures to meet volume and other incentive clauses.

Policies to regulate/eliminate price discounting have been adopted by some states and localities. US states that prohibited the distribution of below-cost coupons to consumers have higher cigarette prices, and thus lower expected cigarette consumption compared to states without a prohibition.^73^ Recently, the states of New Jersey^74^ and New York^75^ implemented coupon redemption bans. However, couponing represents only a small portion of price discounting. Minimum price laws have also been advanced as a potential remedy for discounting practices.^76,77^ These policies set either minimum unit sale prices or minimum wholesale/retail markups.^73^ Almost half of US states have adopted a minimum price law for one or more tobacco products,^78^ but compliance appears to be limited.^79,80^ Two recent studies found that better enforced minimum price laws can have a major impact.^37,81^ However, pack size and other product attributes may also need to be regulated.^71^

Our analysis of marketing expenditures also has implications for Place and Product.^3^ Cigarettes and smokeless tobacco have been sold primarily through mainstream brick-and-mortar retail, especially convenience, drug, and grocery stores, where tobacco companies provide slotting allowances to secure shelf space.^36,82,83^ Part of the promotional allowance expenditures is for branded shelving, complementing the small portion (0.8%) devoted to retail advertising. By limiting available shelf space, these payments may deter competition, particularly from smaller firms and potential new entrants, thereby increasing prices and profits to the major cigarette companies.^17,19,84,85^ In particular, the space made available to potentially less harmful competing products, such as e-cigarettes from independent firms, may decrease. While a significant portion of e-cigarette sales still occurs over the internet and through vape shops, mainstream retail has been gaining market share.^86^ In addition to policies that restrict price discounting payments, policies may be needed to restrict the type of contracts that the tobacco industry can apply to retailers.

A limitation of this analysis is that it is based on mandated reporting of industry expenditures to the FTC, and thus the breakdowns by category may depend on accounting practices of individual firms. In particular, the companies may have incentives to avoid classifying expenditures as advertising in response to the restrictions imposed by the MSA. Our distinction between price reducing and direct advertising may depend on how we classify the component expenditures. For example, expenditures for non-branded items, although not included as a price discounting expenditure, may be viewed as a de facto reduction in consumer price if these items provide benefit to consumers. Further attention should be devoted to understanding the composition of the different types of expenditures.

Although the data only include major firms, these firms account for the vast majority of industry sales.^17,19,87,88^ However, since these data are aggregated to the industry level, the marketing strategies by individual firms cannot be distinguished, e.g., if one firm is a leader in marketing strategies while others follow to maintain market share. In addition, the analysis is for the US, but similar tendencies might be expected in other countries. Price discounting was observed in UK studies before that country banned the practice.^18,40,89^ The WHO Framework Convention on Tobacco Control (WHO FCTC) includes “<u>E</u>nforcing Bans On Tobacco Advertising, Promotion And Sponsorship.” ^90,91^ However, when regulating tobacco marketing, most countries have focused on direct advertising rather than indirect marketing, such price discounting.^3,90,91^

Finally, the FTC reports marketing expenditures only for cigarette and smokeless tobacco, but, price discounting has been documented for flavored cigars^92^ and is featured in cigar ads.^93–95^ Discounts are also utilized by e-cigarette firms,^96^ commonly featured in the tweets of commercial e-cigarette retailers^97,98^ and discounts by vape shops.^99,100^ Further attention should be paid to price discounting for all tobacco products.

## Conclusions

While increasing attention has turned to price discounting behaviors^3,36–38^ and the need to collect data at the retail level,^101^ further research is needed to: (1) better understand industry marketing expenditures for tobacco products other than cigarettes and smokeless tobacco; and (2) evaluate the growing number of state and local policies that aim to increase tobacco prices through non-tax mechanisms. Like industry documents,^11^ the data presented here show that the major cigarette and smokeless tobacco firms view tax increases as a major threat that incentivizes them to discount prices. Such discounts weaken the impact of tax increases and the ability of other firms to gain retail shelf space. Price discounting as a marketing strategy warrants as additional attention. In particular, explicit attention needs to focus on how tobacco companies determine marketing allocations, so that policies can be more effectively directed at counteracting their adverse public health effects.

## Data Availability

Data is publicly available.

## Acknowledgments

This project was funded primarily through National Cancer Institute (NCI) and Food and Drug Administration (FDA) grant U54CA229974. The opinions expressed in this article are the authors’ own and do not reflect the views of the National Institutes of Health, the Department of Health and Human Services, or the United States government. Drs. Levy, Hyland and Cummings also received funding through a grant from the National Cancer Institute (P01CA200512). Dr. Henriksen’s effort was supported by the National Cancer Institute (P01CA225597). We would also like to thank Ken Warner and Jamie Tam for very helpful comments on previous drafts of this paper.

